# Inequalities and policy gaps in maternal health among Empowered Action Group (EAG) States in India: Evidence from Demographic Health Survey

**DOI:** 10.1101/2021.01.15.21249872

**Authors:** Manzoor Ahmad Malik, Saddaf Naaz Akhtar

**Author notes:** **Corresponding author** Saddaf Naaz Akhtar (Ph.D. Scholar), Centre for the Study of Regional Development, School of Social Sciences-III, Jawaharlal Nehru University (JNU), New Delhi, India., Pin Code: 110067, /. **Conflict of Interest statement:** None.

## Abstract

Health inequality in maternal health is one of the serious challenges currently faced by public health experts. Maternal mortality in Empowered Action Group (EAG) states is highest and so are the health inequalities prevalent. We have made a comprehensive attempt to understand maternal health inequality and the risk factors concerning the EAG states in India, using recent data of Demography Health Survey of India (2015-16). Bi-variate, multivariate logistic regression, and concentration indices were used. The study has measured the four outcome variables of maternal health namely antenatal care of at least 4 visits, institutional delivery, contraceptive use, and unmet need. The study revealed that better maternal health is heavily concentrated among the richer households, while the negative concentration index of unmet need clearly reflected the greater demand for higher unmet need among the poor households in the EAG states of India. Challenges of inequalities still persist at large in maternal health, but to achieve better health these inequalities must be reduced. Since inequality mainly affects the poor households due to a lower level of income. Therefore, specific measures must be taken from a demand-side perspective in order to enhance their income and reduce the disparities in the EAG states of India.

## 1. Introduction

Health inequality is an emerging challenge in the public health domain. It is affecting the populations at both the sub-national and national levels [1]. Health Inequalities distress the functioning of the health care system and its utilization through multiple factors, linked with the well-being of individuals [2]. Inequality in health reflects the difference in health status among individuals, groups, or geographical locations within or across countries [3]. Thus, the distribution of health in the population is a key factor to determine the public health performance [4]. Health inequality is particularly predominant in vulnerable groups of the population which are at greater risk like women, children, and the elderly [5]. Health inequalities can be multiple depending on the type of health care system taken into account like inequalities in maternal, child, elderly, or adult health. Health inequalities become important when they have resulted in greater morbidity and mortality outcomes.

Health inequality in maternal health is one of the serious challenges currently faced by public health experts. It implies serious challenges since inequality in maternal health is directly linked with child health and maternal death [6]. Maternal health inequality enhances the risk of maternal conditions, which increase the likelihood of disability and child growth [7]. Inequalities in maternal health have greater consequences on maternal and child health. These inequities not only enhance mortality and morbidity levels among women, but they also have an impact on the child’s health as well. Further, the increase the health burden through economic costs, and welfare distribution particularly in terms of yielding better health outcomes affect the overall public health system [8], which can be lowered while reducing the maternal health inequality in order to lower the risk of maternal death and morbidity [9]. Equity has now become central to health outcomes [10], but health inequality still persists at large due to the factors like access, awareness, and affordability apart from socio-economic and geographical differentials. Therefore, investing in maternal health is an important way to improve the overall health systems of the country and reduce health inequalities [11].

### 1.1 Theoretical construct

Inequality in health care services is gaining global attention due to poor health conditions especially among the vulnerable populations. Multiple studies have examined the determinants of health care inequalities and the burden they impose on levels of human development [12]. There are numerous factors that affect the level of inequality in maternal health, but some of the detrimental factors include antenatal care visits, institutional deliveries family planning use, and demand for unmet need. Studies have significantly highlighted the role of antenatal care in lowering the risk of maternal deaths [13]. Similarly, the role of institutional deliveries and better family planning use is also key to lowering the risk of maternal and child deaths. Inequality in maternal health not only result due to these institutional factors, but socio-economic and demographic factors play an important role in lowering the risk, such as age education, working status, and religion, apart from literacy, gender bias, socio-political environment, and quality of care [14–16]. Inequalities in maternal health also result due to lack of access, awareness, and availability to health care systems [10]. There is also emerging evidence of the link between poverty and maternal deaths in low- and middle-income countries [17]. Cross country studies in developing countries have shown that deaths due to maternal health consequences have consistently increased due to the rise in poverty. Furthermore, gender inequality and women’s low social status and disempowerment significantly impact maternal health and demand for maternal healthcare services [18].

### 1.2 Maternal health in Indian settings

India is one of the countries with greater inequalities in health care [19]. Although India’s maternal and child mortality rate has declined over the last two decades (SRS Reports), but health differentials are still very high at the regional level. India is not too far in the list of countries with the slowest improvement in terms of maternal health. Despite implementing numerous programs and policies, maternal health inequality is still higher in India. While only little achievement could be made from the MDG perspective which was less conclusive. Earlier studies have shown that large inequalities exist due to socio-economic and demographic factors [20]. But there are other factors like literacy, awareness, and regional differences attributed to this rising health inequality in India. According to a study conducted by [21] economic status is one of the major factors, which result in a higher level of inequality in India. Whereas another reason is the multiple deprivations, which increases the risk of health care utilization [22]. Similarly, factors like poverty and lack of access result in persistent health inequalities [23,24]. Inequalities also persist highly due to regional and rural-urban differentials [20,25].

India is now aiming for achieving the target of Sustainable Development Goals, through National Health Policy (NHP), that aims to bring down maternal mortality ratio to 100 by the year 2020 [26] but to achieve these targets it is necessary to understand the level of inequality that affecting the maternal level outcomes and our study is one of such attempts. Maternal mortality in Empowered Action Group (EAG) states is highest and so is the health inequalities prevalent in these states [27]. Accounting 48% of India’s total population EAG states are at greater risk due to the level of health disparities [28]. Although numerous attempts have been carried out earlier to reduce the disparities across health sectors. But despite these attempts and policy interventions, maternal and reproductive health is unacceptably emerging as a challenge in India. Thus, the present study will make a comprehensive attempt to understand maternal health inequality and the risk factors with respect to the EAG states in India.

## 2. Materials and Methods

### 2.1 Materials

Data for the study was used from the fourth round of Demography health Survey of India also called as National Family Household Survey (NFHS). The survey included all regions of India, but our focus was especially on the EAG states only. National Family Health Survey is a large-scale, multi-round survey conducted in a representative sample of households throughout India. NFHS-4 is based on a sample of 699,686 women in 601,509 households with a response rate of 98%. The survey included 425,563 households from rural areas and 175,946 households from urban areas. The sample size for NFHS-4 is based on a two-stage sample design. NFHS-4 data provides up to district level information on socio-demographic and health indicators like fertility, family planning, infant and child morbidity and mortality, maternal and reproductive health, nutritional status of women and children, and the quality of the health services. Our study mainly focuses on EAG states with an estimated sample size of 353480.

### 2.2 Methods

Bivariate analysis including prevalence and proportions were used to examine the distribution of socio-demographic, economic, and maternal health outcomes. The factors associated with dependent variables were examined using first the logistic regression and then probabilities were computed to understand the dynamics. There are a number of methods used to examine the health disparities among households and their health care utilization like equity gaps, equity ratios, concentration curve and concentration indices [29]. We used concentration indices to measure the health inequality. Income-related maternal health inequality was assessed by plotting the cumulative proportion of health across individuals ranked from poorest to richest. All analyses were conducted using STATA 14.0.

## 3. Results

Table 1 shows the sample distribution of maternal health indicators by Empowered Action Groups (EAG) states of India for the currently married women. There are 353480 samples of currently married women in the EAG states of India. The highest receiving percentage of ANC 4+ was found in Odisha (63%) followed by Chhattisgarh (59.55 %) and Rajasthan (38.89 %), but the lowest receiving percentage of ANC 4+ was found in Uttar Pradesh (26.52%) followed by Uttarakhand (4.89 %). The use of contraceptive by women in Rajasthan (57.7%) is highest in comparison to the other EAG states and lowest in Bihar (24.1%). The highest institutional birth is found in Odisha (69.47%) whereas the lowest is found in Jharkhand (42%). The unmet need for family planning is found to be lowest in Madhya Pradesh (12.14%) and highest in Bihar (21.15%). Accessing health care problems are highly faced by women in Odisha (86.79%) followed by Bihar (83.66%), Chhattisgarh (82.29%) and Jharkhand (82.27%).

**Table 1.** Percentage of currently married women using maternal health indicators for Empowered-Action-Group (EAG) states of India from National Family Health Survey (Round-IV).

| ***EAG States*** | ***ANC 4+*** | ***C-Use*** | ***Institutional***<br>***Delivery*** | ***Unmet***<br>***Need*** | ***Sample***<br>***Size*** |
| --- | --- | --- | --- | --- | --- |
| *Bihar* | 14.81 | 24.1 | 48.61 | 21.15 | 45,812 |
| *Chhattisgarh* | 59.55 | 57.7 | 57.25 | 11.11 | 25,172 |
| *Jharkhand* | 30.44 | 40.4 | 42.54 | 18.37 | 29,046 |
| *Madhya*<br>*Pradesh* | 37.53 | 51.4 | 69.47 | 12.14 | 62,803 |
| *Odisha* | 63.19 | 57.3 | 76.08 | 13.61 | 33,721 |
| *Rajasthan* | 38.89 | 59.7 | 63.8 | 12.31 | 41,965 |
| *Uttar Pradesh* | 26.52 | 45.5 | 44.91 | 18.05 | 97,661 |
| *Uttarakhand* | 32.05 | 53.4 | 45.49 | 15.54 | 17,300 |
| ***Total*** | 31.3 | 45.3 | 53.82 | 16.34 | 3,53,480 |

*Source: Fourth round of National Family Health Survey (NFHS-IV) of India (2015-16).*

Our study measured the four outcome variables of maternal health namely antenatal care of at least 4 visits, institutional delivery, contraceptive use, and unmet need. The basic socio-demographic characteristics with percentages of the respondents in EAG states were presented in Table 2.

**Table 2.** Showing the percentage of women currently married by background characteristics using maternal health indicators for Empowered-Action-Group (EAG) of India from National Family Health Survey (Round-IV).

| **Background Characteristics** | **ANC 4+** | **C-Use** | **Institutional Delivery** | **Unmet Need** |
| --- | --- | --- | --- | --- |
| **Age Groups** |  |  |  |  |
| *Below 20* | 29.51 | 8.6 | 80.64 | 25.58 |
| *20-30* | 33.25 | 31.7 | 76.47 | 24.09 |
| *30-40* | 28.56 | 57.6 | 68.61 | 14.02 |
| *40+* | 16.17 | 55.5 | 50.82 | 6.22 |
| **Place of Residence** |  |  |  |  |
| *Urban* | 48.39 | 53.74 | 82.84 | 13.95 |
| *Rural* | 27.1 | 42.85 | 71.36 | 17.03 |
| **Education** |  |  |  |  |
| *No Education* | 18.28 | 45.61 | 60.75 | 14.29 |
| *Primary* | 29.36 | 47.95 | 72.54 | 15.7 |
| *Secondary* | 40.01 | 43.83 | 83.63 | 18.62 |
| *Higher* | 58.92 | 44.77 | 93.79 | 19.97 |
| **Religion** |  |  |  |  |
| *Hindu* | 31.91 | 47.11 | 75.44 | 15.65 |
| *Muslim* | 27.11 | 32.3 | 63.77 | 21.38 |
| *Other* | 37.02 | 44.31 | 66.77 | 16.04 |
| **Caste** |  |  |  |  |
| *ST* | 25.56 | 44.16 | 70.72 | 16.8 |
| *SC* | 34.52 | 44.44 | 64.77 | 14.64 |
| *OBC* | 29.62 | 44.85 | 74.23 | 16.51 |
| *OTH* | 41.41 | 48.86 | 81.35 | 16.37 |
| **Wealth Index** |  |  |  |  |
| *Poorest* | 17.42 | 36.11 | 60.56 | 18.93 |
| *Poor* | 27.85 | 44.25 | 74.46 | 16.39 |
| *Middle* | 36.31 | 48.79 | 80.42 | 15.23 |
| *Richer* | 45.49 | 51.64 | 85.03 | 15.01 |
| *Richest* | 62.28 | 57.37 | 92.77 | 13.04 |
| **Work Status** |  |  |  |  |
| *Non-Working* | 33.28 | 44.47 | 75.57 | 17.28 |
| *Working* | 28.41 | 56.61 | 66.04 | 12.23 |
| **Total** | 24.73 | 53.03 | 73.7 | 16.24 |

Source: Fourth round of National Family Health Survey (NFHS-IV) of India (2015-16).

The results in Table 3 show the predicted probabilities for the outcome variables based on wealth and residence. The resulted show that the average predicted probability of women currently married using ANC 4+ in the rural areas is 5% higher than that of urban areas for Empowered-Action-Group (EAG) states of India. While as it was found to be lower among the urban women in case of contraceptive use institutional delivery and Unmet need as shown in Table 3.

**Table 3.** Probability of currently married women Residence and Wealth Index using maternal health indicators for Empowered-Action-Group (EAG) of India from National Family Health Survey (Round-IV).

| *Background*<br>*Characteristics* | *ANC 4+* | *C-Use* | *Institutional*<br>*Delivery* | *Unmet Need* |
| --- | --- | --- | --- | --- |
|  | Margins with [Confidence Interval (%)] |  |  |  |
| **Place of**<br>**Residence** |  |  |  |  |
| *Rural* | 0.33 [0.33,0.33] | 0.36 [0.36,0.36] | 0.75 [0.75,0.75] | 0.11 [0.11,0.11] |
| *Urban* | 0.37 [0.37,0.38] | 0.34 [0.34,0.35] | 0.73 [0.73,0.74] | 0.10 [0.10,0.10] |
| **Wealth Index** |  |  |  |  |
| *Poorest* | 0.21 [0.21,0.22] | 0.30 [0.30,0.30] | 0.61 [0.61,0.62] | 0.13 [0.12,0.13] |
| *Poorer* | 0.31 [0.30,0.31] | 0.35 [0.35,0.35] | 0.75 [0.75,0.76] | 0.11 [0.11,0.11] |
| *Middle* | 0.38 [0.37,0.39] | 0.37 [0.37,0.38] | 0.81 [0.80,0.81] | 0.10 [0.10,0.10] |
| *Richer* | 0.46 [0.45,0.47] | 0.39 [0.38,0.39] | 0.86 [0.85,0.86] | 0.10 [0.10,0.11] |
| *Richest* | 0.61 [0.60,0.62] | 0.42 [0.41,0.42] | 0.93 [0.93,0.94] | 0.09 [0.09,0.10] |

Source: Fourth round of National Family Health Survey (NFHS-IV) of India (2015-16).

Similarly, while calculating the predicted probability based on logistic regression, we found that poorest has the lowest probability of using any maternal health indicator except unmet need which is highest them. It can be found from Table 3 that the probability of using contraceptive is 30% only to that poorer which is around 35%. This clearly should be the higher percentage of probabilities in terms of demand for unmet need. Table 4 presents the results computed from concentration indices computed for the outcome variables against the wealth-income. The results clearly indicate that better maternal health is heavily concentrated among the richer households, while the negative concentration index of unmet need clearly reflects the greater demand for higher unmet need among the poor households in the EAG states.

**Table 4.** Concentration indices of currently married women using maternal health indicators for Empowered-Action-Group (EAG) of India from National Family Health Survey (Round-IV).

| *Maternal health indicators* | *Index values* | *Robust Std. Error* |
| --- | --- | --- |
| *ANC 4+* | 0.25 | 0.004 |
| *C-Use* | 0.07 | 0.002 |
| *Institutional Birth* | 0.08 | 0.002 |
| *Unmet Need* | -0.09 | 0.004 |

*Source: Fourth round of National Family Health Survey (NFHS-IV) of India (2015-16).*

Fig. 1 to Fig. 4 shows the concentration curves of outcome variables by wealth Index. These curves clearly show the greater levels of inequality in the maternal health among the poorest households in the EAG states.

**Fig. 1.**
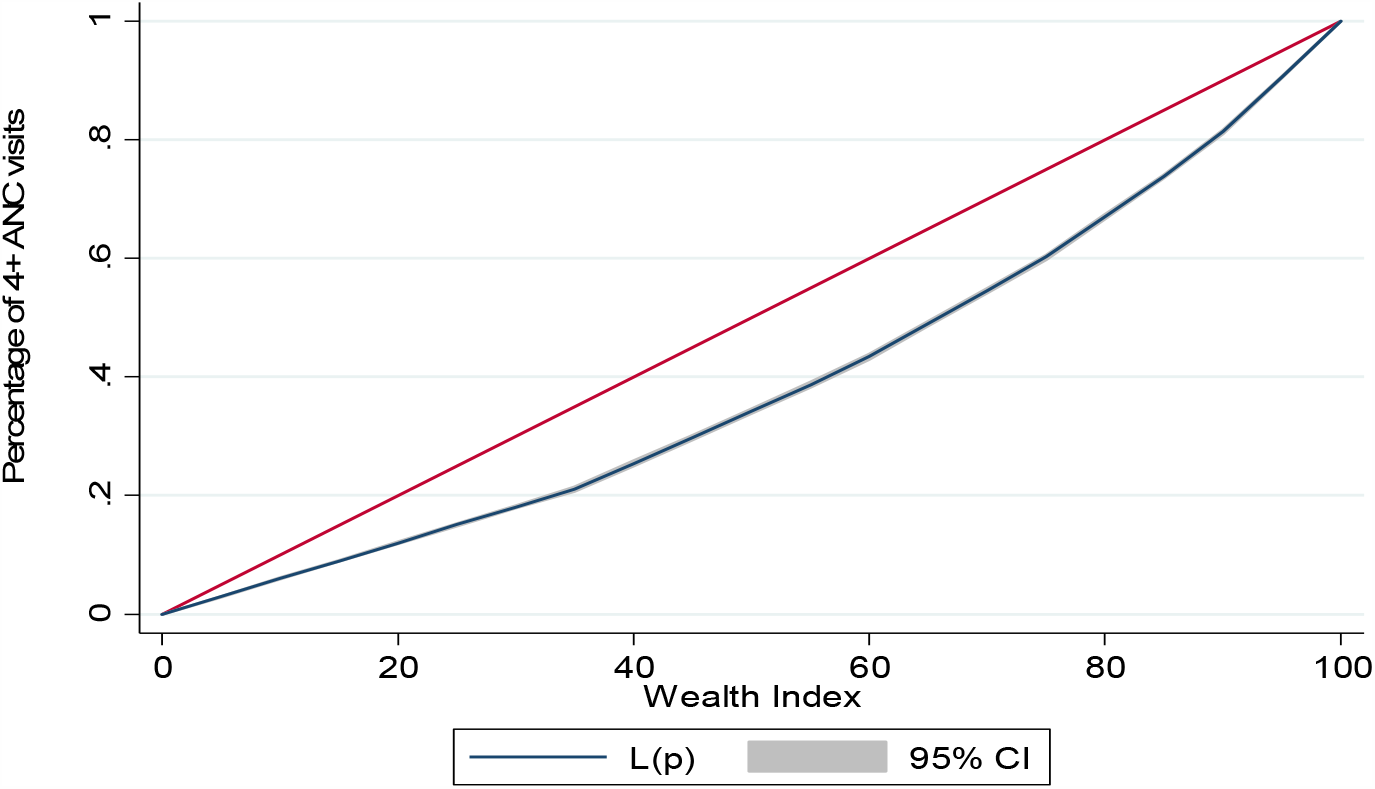
Concentration curve of ANC 4+ by wealth index of EAG states’ households of India from National Family Health Survey (Round-IV).

**Fig. 2.**
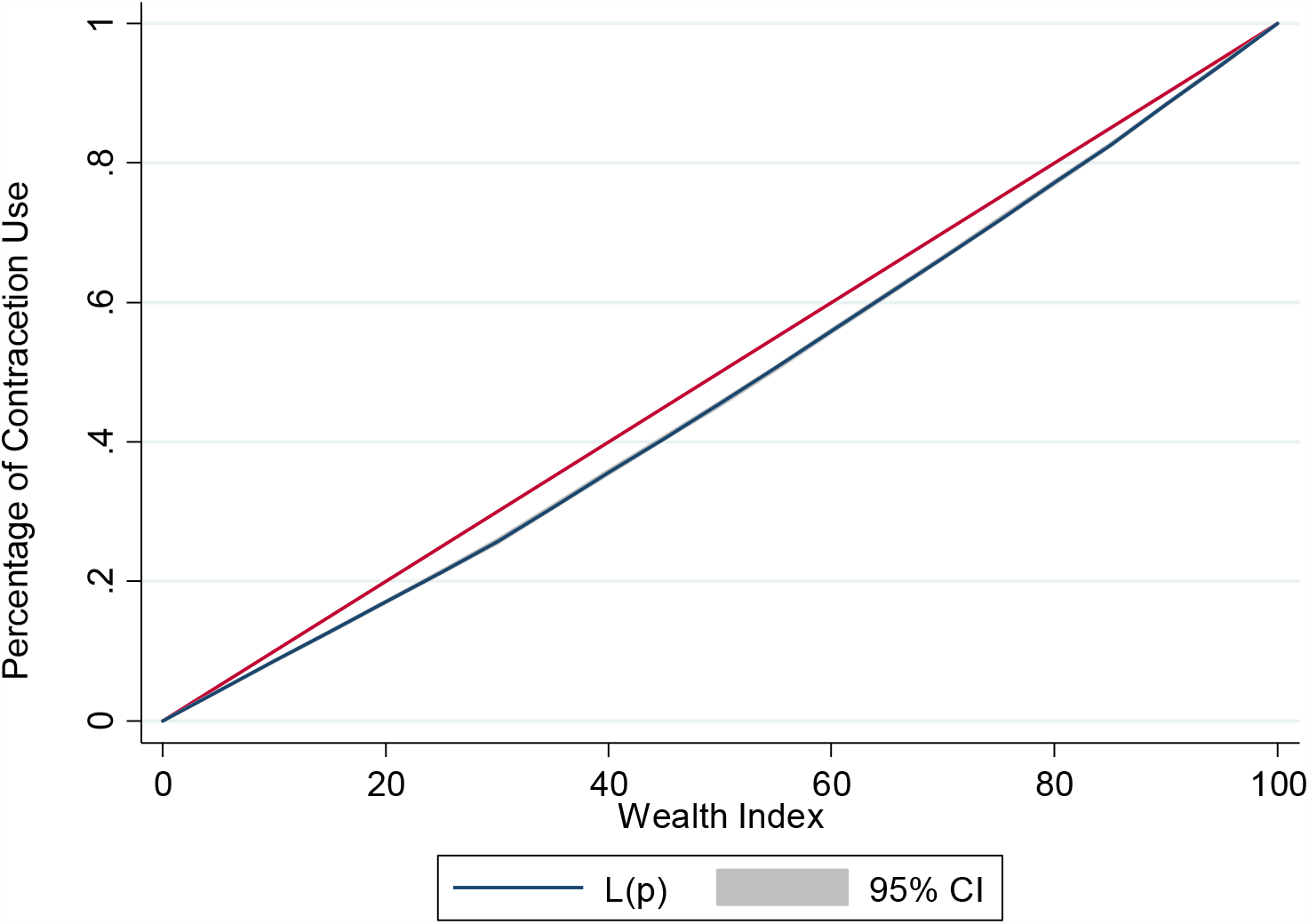
Concentration curve of contraceptive use by wealth index of EAG states’ households of India from National Family Health Survey (Round-IV).

**Fig. 3.**
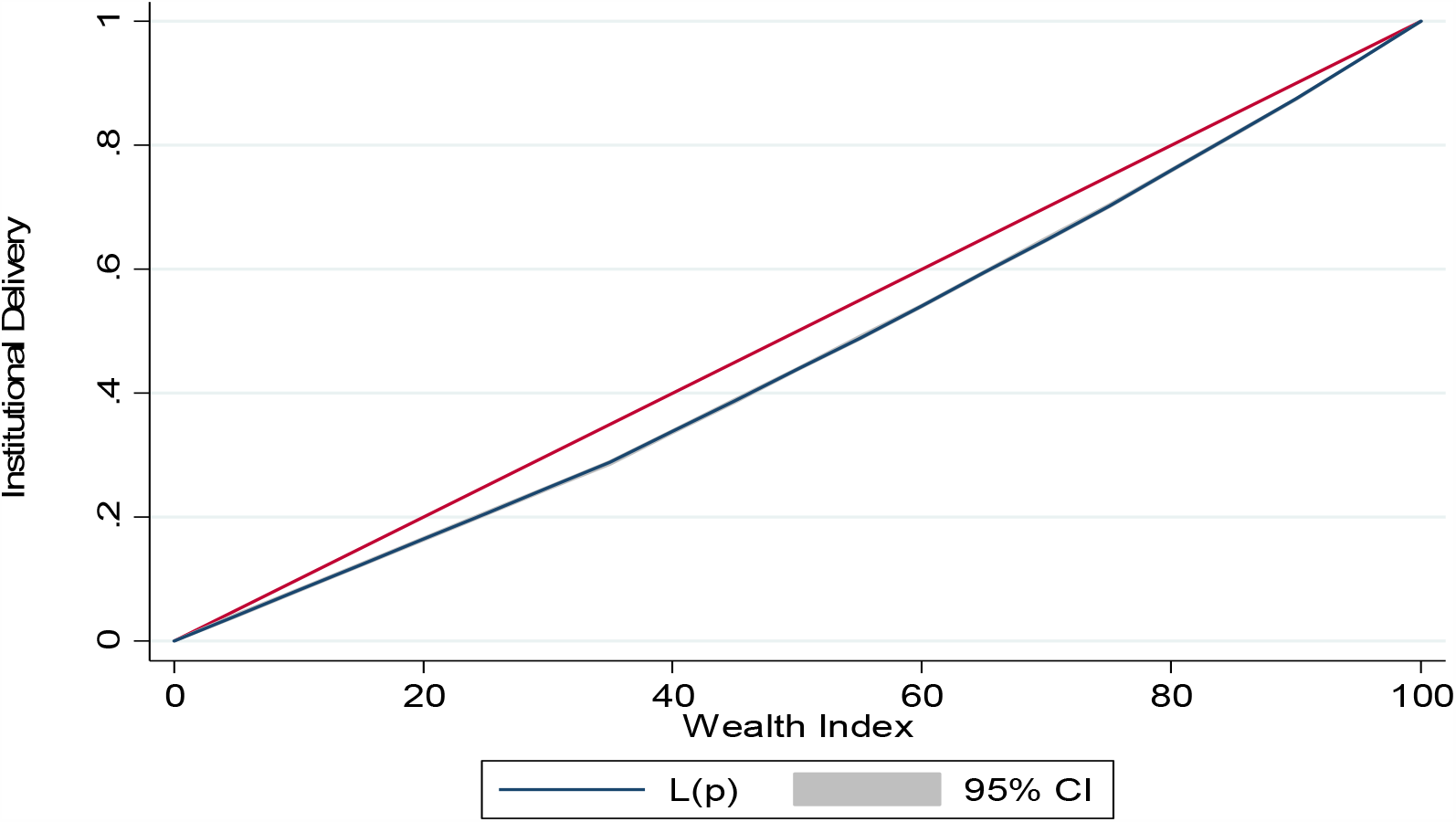
Concentration curve of institutional delivery by wealth index of EAG states’ households of India from National Family Health Survey (Round-IV).

**Fig. 4.**
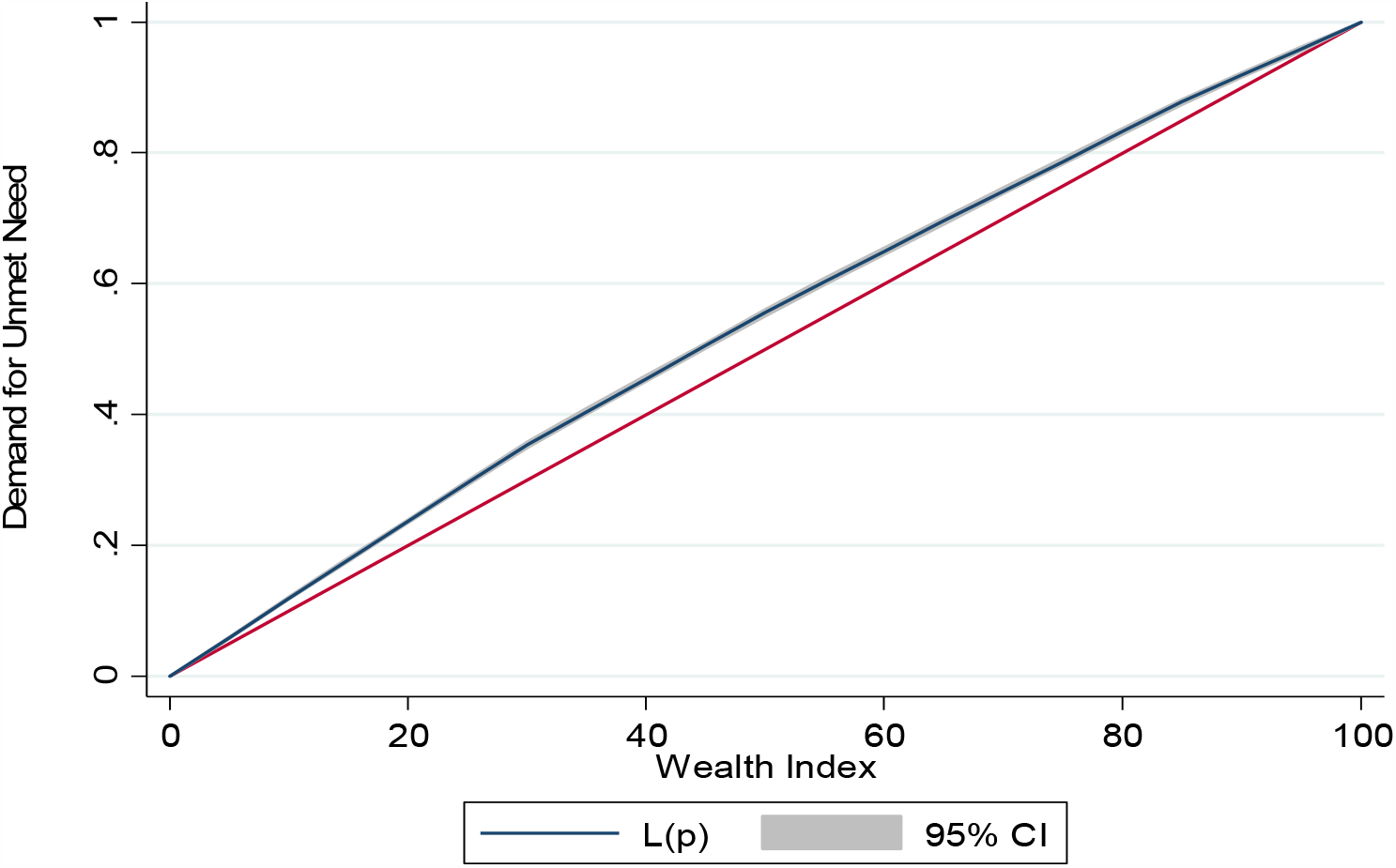
Concentration curve of unmet need by wealth index of EAG states’ households of India from National Family Health Survey (Round-IV).

## 4. Discussion

Health plays a significant role in driving the economy. A good and equitable health system lessens the economic burden and minimizes the risk of mortality and morbidity outcomes [30]. Studies measuring health disparities have grown on a scale over time to underlie the factors that help in measuring the disparities and determine the factors which help us to measure the health inequalities [31–33]. Similarly, various studies have tried to examine the inequality in maternal health care due to various factors of being socio-economic and demographic in nature. It is clear from our results that within region inequality is highly persistent in India as shown by the varying maternal health indicators across the regions (NFHS-4). Our findings clearly suggest that the largest gap results in coverage between the richest and poorest, due to their socio-economic differentials. These inequalities can be clearly seen in the case of ANC Visits, institutional delivery, and contraceptive use well supported by previous other studies as well [34,35]. Thus, the inequality gap has not reduced over time. Inequality in maternal health care is a critical issue and must be addressed. There are several factors contributing to its persistent prevalence among poor households as found in our study. Our study is consistent with the earlier studies that reflected that economic marginalization is a key factor to reflect the greater level of inequality in maternal health among poor households [21].

The results in the study showed that there is a substantial inequality present among all the outcome variables between poor and rich households in the EAG states. Poor households are at more risk since 50% of the richest group is benefiting from the services than the poorest households. This reflects the vulnerability among poor people to benefit from maternal health care services. Previous studies in this context have almost reflected similar outcomes like [36] who found that inequality in health facility delivery is lowest among the poor. While like the other reason reflected in this persistent inequality is the higher cost, which keeps women from poor households at bay to reap out the benefits from the maternal health care [37,38]. Similarly, inequality in ANC services among the poor is due to poor coverage and access at which involves costs that are less affordable to poor households [39]. Although the study could not emphasize on the access and affordability, the determinant factors that result in affordability is mainly the lack of access. Unmet need is one of the keys. Thus, it depends upon the kind of services providers and the lack of communication between the households especially poor. Our findings are in line with what has been emphasized by the earlier studies in reasons for access and affordability as a key concern in terms of rising inequalities of health care outcomes. The policies and programs must aim from strengthening the utilization services while improving accessibility and awareness especially among the poor women households. Both the supply and demand-side factors must be targeted coincide in order to attain the equity in maternal health coverage. Poor households are likely at greater risk from profiting the health care services provided. While there is an urgent need to reduce the inequalities in maternal health. Adequate access to factors like antenatal care, institutional delivery, family planning use, and other numerous maternal health indicators must be provided at large. Since full ANC is essential in lowering the risk of maternal health [40], it should be prioritized among the poor. Similarly, Institutional based delivery helps to reduce maternal mortality and morbidity [13], which must be taken care of in order to reduce inequality and provide an equal distribution of health. While issues of unmet need must be addressed in order to avert the challenges of family planning. Furthermore, access can also be pivotal in lowering the risk of inequality, but that must be provided with the domain especially of poor households. Because health care utilization can be optimal provided better access [29].

## 5. Conclusions and recommendation

Thus, to sum up, although the challenges of inequalities persist at large in maternal health, but to achieve better health, these inequalities must be reduced. At the same time, it is clear from the above that inequality mainly affects the poor households due to lower levels of income. Inadequate utilization of healthcare services has been observed in the EAG states, especially among low-income families, which revealed that there is a need to improve the quality of public healthcare services. Due to lack of knowledge, religious objections and access to healthcare services in Bihar, the use of contraception is lowest, thus, there is a need to encourage and widespread the knowledge about the use of contraception. Also, it is essential to strengthening the health infrastructure in Jharkhand to increase the institutional delivery and this can play a crucial role in reducing maternal health consequences. The State Government of EAG states should establish an equitable approach to healthcare services for everyone. Therefore, specific policy must be taken from a demand-side perspective to enhance their income and reduce the health disparities in the EAG states of India.

## Data Availability

Data for the study was used from the fourth round of Demography health Survey of India also called as National Family Household Survey (NFHS). The data is available at https://dhsprogram.com/methodology/survey/survey-display-355.cfm

## Acknowledgement

None.

